# Need of care in interpreting Google Trends-based COVID-19 infodemiological study results: potential risk of false-positivity

**DOI:** 10.1101/2020.12.30.20249066

**Authors:** Kenichiro Sato, Tatsuo Mano, Atsushi Iwata, Tatsushi Toda

## Abstract

**Objective:** Google Trends (GT) is being used as an epidemiological tool to study coronavirus disease (COVID-19) by identifying keywords in search trends that are predictive for the COVID-19 epidemiological burden. However, many of the earlier GT-based studies include potential statistical fallacies by measuring the correlation between non-stationary time sequences without adjusting for multiple comparisons or the confounding of media coverage, leading to concerns about the increased risk of obtaining false-positive results. In this study, we aimed to apply statistically more favorable methods to validate the earlier GT-based COVID-19 study results.

**Methods:** We extracted the relative GT search volume for keywords associated with COVID-19 symptoms, and evaluated their Granger-causality to weekly COVID-19 positivity in eight English-speaking countries and Japan. In addition, the impact of media coverage on keywords with significant Granger-causality was further evaluated using Japanese regional data.

**Results:** Our Granger causality-based approach largely decreased (by up to approximately one-third) the number of keywords identified as having a significant temporal relationship with the COVID-19 trend when compared to those identified by the Pearson correlation-based approach. “Sense of smell” and “loss of smell” were the most reliable GT keywords across all the evaluated countries; however, when adjusted with their media coverage, these keyword trends did not Granger-cause the COVID-19 positivity trends (in Japan).

**Conclusion:** Our results suggest that some of the search keywords reported as candidate predictive measures in earlier GT-based COVID-19 studies may potentially be unreliable; therefore, caution is necessary when interpreting published GT-based study results.

## Introduction

Google Trends (GT) is a publicly available source of online Google search trafficking data (https://trends.google.co.jp/trends), which allows users to visualize changes in time series related to the general public’s online interest in certain keywords. It is used as one of the “infodemiology” tools [1] to study epidemiological trends of certain disease outbreaks such as the Middle East Respiratory Syndrome epidemic and the Ebola outbreak [1]. As for coronavirus disease (COVID-19) that became a worldwide pandemic in early 2020 [2], the potential use of GT to predict COVID-19 cases or deaths has been reported with regard to GT trends and keyword searches of “COVID-19” [3, 4] or any of its symptoms, including chest pain, anosmia, dysgeusia, headache, shortness of breath, etc. [5-7] within the initial months following the outbreak [4-9].

In many earlier studies analyzing GT trend data as an epidemiological tool, with a few exceptions [10-12], analytical fallacies were of concern. First, Pearson (or Spearman) correlation is often applied to assess the correlation between the time-series trends of COVID-19 cases/deaths and GT trends in symptom keywords without confirming the stationarity of these time series, which is sometimes critically inappropriate in the context of time-series analyses because it can increase the likelihood of obtaining spurious correlations. Second, the Pearson/Spearman correlation tests were repeated for each of the included symptom keywords (e.g., fever, cough, pneumonia, anosmia, sore throat, headache, etc. [7]) without adequate adjustment for multiple comparisons, which would also increase the risk of false-positive results. Third, because COVID-19 and its symptoms have attracted intensive attention worldwide, the influence of media coverage on GT symptom keywords is inevitable [9, 13, 14], which has hardly been adjusted in a statistically favorable manner.

Based on the above analytical concerns for earlier studies, by using the vector autoregression (VAR) model [10-12] in this study, we aim to identify statistically more reliable symptom keywords for which GT trends may be used as a predictive measure for future COVID-19 positivity trends, and to validate the earlier study results.

## Methods

### Extracting Google Trends and COVID-19 data

All the following data handling and analyses were performed using R 3.5.2 (R Foundation for Statistical Computing, Vienna, Austria). A statistical level of less than 0.05 is considered significant if not stated otherwise. COVID-19 data and Google Trends (GT) data were separately analyzed in nine different regions: Japan (JP) and eight English-speaking countries, namely, Australia (AU), Canada (CA), Great Britain (GB), Ireland (IE), India (IN), Singapore (SG), United States (US), and South Africa (ZA).

The three-year (October 1, 2017–October 25, 2020) time series GT trend data for keywords of symptoms that may be related to COVID-19 was queried using R package *gtrendsR* [15]. Individual queries were separately conducted for each keyword in all nine regions. Search keywords were defined as listed in Table 1: 54 English keywords were used for search in eight English-speaking country regions, and the corresponding 60 Japanese keywords (as listed in S1 Table) were used for searches in the Japan region. The obtained data were the weekly relative search volume for each keyword, of which the maximum value during the included period was normalized to 100%. For the timings when the relative search volume was 1% or less, we imputed them as 0%.

**Table 1.** Included English and Japanese keywords search for Google Trends.

| English keywords searched in Google Trends |  |  |
| --- | --- | --- |
| malaise | fatigue | tired |
| anorexia | diarrhea | constipation |
| abdominal pain | stomach ache | nausea |
| chest pain | dyspnea | vomiting |
| shortness of breath | short of breath | pneumonia |
| cough | sputum | rhinitis |
| runny nose | nasal discharge | stuffy nose |
| sneeze | sore throat | throat pain |
| fever | chills | cold |
| sense of smell | loss of smell | anosmia |
| sense of taste | loss of taste | dysgeusia |
| hair loss | loss of hair | bald |
| myalgia | muscle pain | body aches |
| arthralgia | joint pain | pain |
| eye pain | sore | congestion |
| headache | memory loss | confusion |
| vertigo | dizziness | dizzy |
| insomnia | anxiety | numbness |

Search keywords were arbitrarily defined: 54 English keywords were used for search in 8
English-speaking country regions, and the corresponding 60 Japanese keywords (as listed in S1 Table)
were used for search in the Japanese region.

For COVID-19 data on serial daily number of positive cases from January 22, 2020, we downloaded data from the web database (https://data.humdata.org/dataset/novel-coronavirus-2019-ncov-cases, accessed on October 30, 2020) provided by the United Nations Office for Coordination of Humanitarian Affairs. Since we did not include the number of positive cases from mainland China, we imputed the number of COVID-19 cases before January 22, 2020 as zero (even for 2017–2019). The COVID-19 daily case data were converted to weekly serial data, in reference to the above GT weekly trend data.

### Preprocessing and analysis

The keyword weekly trend data were further processed as shown in Fig 1. Fig 1A (uppermost row) is the three-year original GT time series for “chest pain” in the United States region. The sequence was processed using R package *stats* to remove seasonality (one-year level) and the general trend from the original series, and the remaining random series (Fig 1A, lowermost row) was used as the keyword trend data to analyze [10]. Then, the obtained series were evaluated with an augmented Dickey–Fuller (ADF) test using R package *tseries* [16] to examine whether the sequence was stationary (Fig 1B). If the series was not considered stationary, the sequence was further differenced so that the differenced series became stationary (as confirmed by the ADF test again).

**Fig 1.**
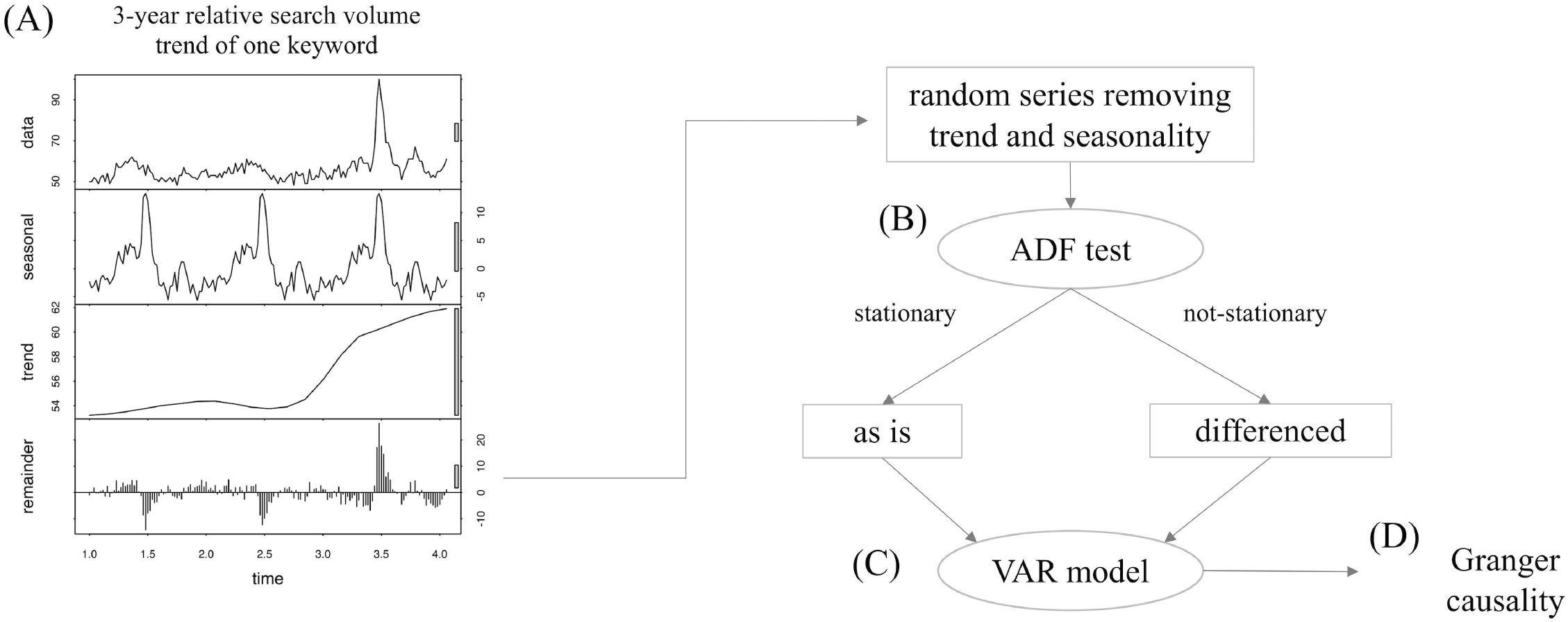
Outline of preprocessing flow. The sequence was processed to remove seasonality (1-year level) and general trend from the original series, and the remaining random series (A, lowermost row) was used as the keyword trend data. Then, the obtained series were evaluated with an ADF test to examine its stationarity (B). Next, the temporal relationship between the processed sequence of each single keyword and the COVID-19 weekly positivity data was analyzed using the VAR model (C). Then we assessed whether the keyword trend Granger-causes COVID-19 positivity trends (D).

Next, the temporal relationship between the processed sequence for each keyword and the COVID-19 weekly positivity data was analyzed with the VAR model [10, 11] (Fig 1C), using R package *vars* [17]. Since the COVID-19 weekly positivity trend data was actually not stationary by itself, its difference sequence was imputed to the VAR analysis. The adequate lag was determined from the lag order range of 1-4, 1-6, or 1-8, based on the Akaike’s information criterion. Then, using the obtained VAR model, we assessed whether the keyword’s trend Granger-caused the COVID-19 positivity trends [10, 11] (Fig 1D). This implied that the change in the keyword trend could have the potential to practically predict the near-future change in the COVID-19 positivity trend. The causality here was merely a statistical one and did not require true causal mechanisms between the two trends. One p-value was obtained for the Granger-causality of one keyword to the COVID-19 trend and the Granger-causality analysis was performed for all the keywords. We adjusted multiple testing using the Benjamini-Hochberg (BH) method [18] within the country-wise groups. The BH method regulates the false discovery rate (FDR), which has a smaller risk of false-positivity than the raw p-value and is more powerful than the most stringent Bonferroni method.

In addition, as a reference, we also calculated the Pearson correlation between the raw GT keyword trends and the COVID-19 weekly positivity trends, as in the earlier GT-based COVID-19 studies. Pearson’s p-values were similarly adjusted with the BH method.

### Incorporating media coverage trends

We then evaluated the media coverage of the obtained GT keywords with a statistically reliable temporal relationship with the COVID-19 weekly positivity trend. Due to the shortness of available data, we could only analyze the media coverage trend of those keywords in the Japan region. We reviewed *Nikkei Telecom* (http://telecom.nikkei.co.jp), a large Japanese database covering newspapers, TV news, Internet news, and general magazines published in Japan, to measure the weekly number of published articles in which the title/abstract/manuscript included the identified Japanese keyword. Specialized magazines were excluded from the reviewed publication review because they might have less exposure to the general population. The obtained time series of the weekly count of articles containing the keyword was used as the media coverage trend in Japan. Then, we again evaluated whether the identified GT keyword trend still Granger-caused the COVID-19 weekly positivity, even when adjusted with the simultaneous media coverage trend of the keyword. This partial Granger-causality analysis was performed using the R package *FIAR* [19].

### Ethics

This study was approved by the University of Tokyo Graduate School of Medicine Institutional Ethics Committee (ID: 11628-(3)). Informed consent was not required because the data were publicly distributed. The study was conducted in accordance with the ethical standards laid out in the Declaration of Helsinki, 1964.

## Results

### General COVID-19 related trends

During the three-year period from October 1, 2017 to October 25, 2020, different countries experienced different timings in their COVID weekly positivity trends and the related GT search trends. Fig 2 shows weekly trends of each country (from upper-left to lower-right in alphabetical order by country code). The solid lines show the weekly COVID-19 positivity trends while the dotted lines denote GT search volume trends for the “COVID” keyword in each region (or its corresponding Japanese keyword in Japan). Both trends are plotted in a normalized manner so that the maximum value of each trend within the reviewed period becomes 100%. Briefly, as of late October 2020, for both the COVID-19 weekly positivity trend and the COVID search volume trends, Australia (AU), Japan (JP), and the United States (US) experienced their first and second waves (i.e., large positive peaks), while Canada (CA), Great Britain (GB), and Ireland (IE) are currently experiencing their second wave. Meanwhile, although India (IN) and South Africa (ZA) experienced delayed first waves of weekly COVID-19 positivity compared to other countries, search volume trends showed the first wave surge, the timing of which was similar to that of the other countries.

**Fig 2.**
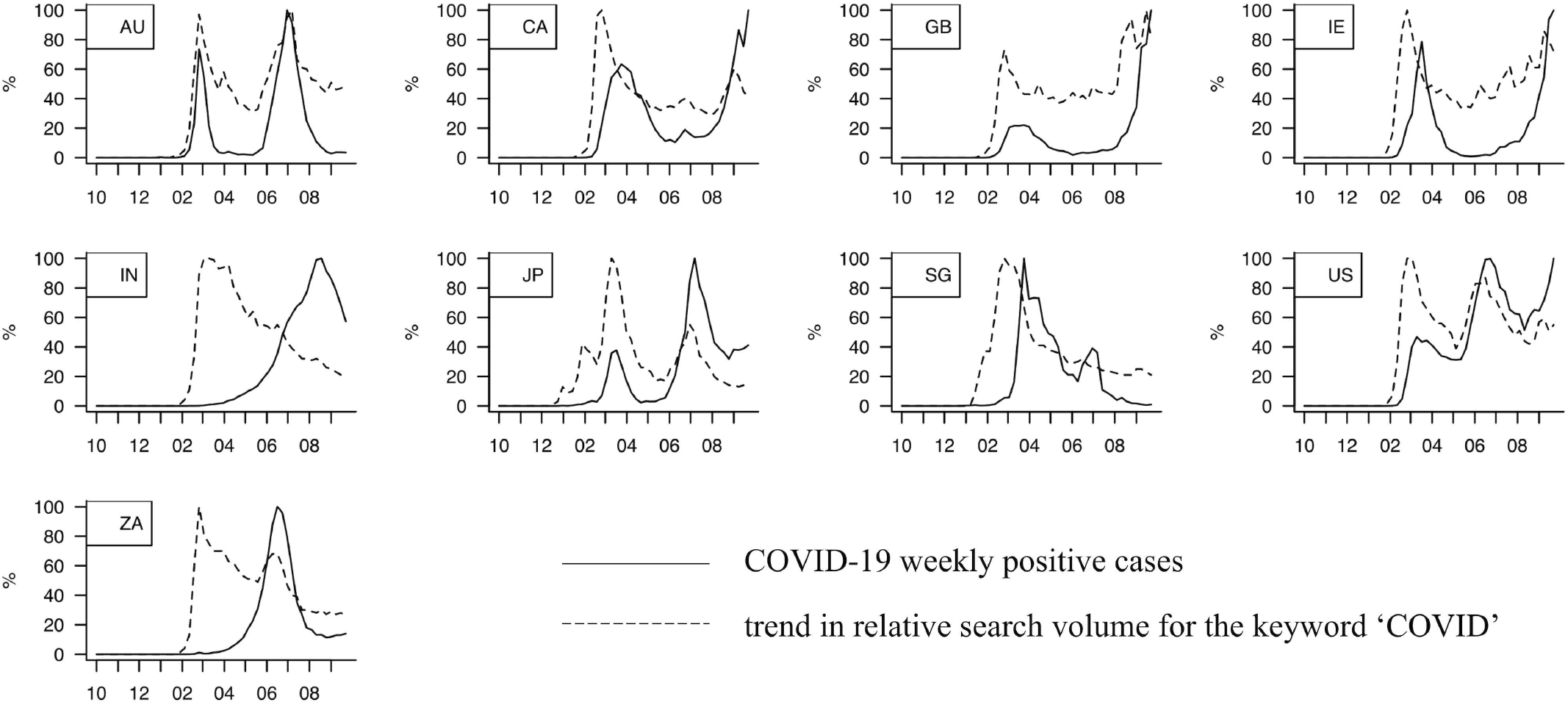
COVID-19 weekly positivity trends and related GT search volume trends for ‘COVID’ in each region. Figures show weekly trends of each country (from upper-left to lower-right in alphabetical order of country name), where the solid lines show COVID-19 positivity trends while the dotted lines denote GT search volume trends for ‘COVID’ word (or its corresponding Japanese word) in each region. Both trends are plotted in a normalized manner so that the maximum value within the period becomes 100%. X-axis in months since October 2019 to September 2020.

### VAR model in comparison with Pearson correlation

Next, we conducted a VAR model analysis. Table 2 summarizes the number of keywords of which GT trends had significant (p-value or FDR < 0.05) temporal relationships with the COVID-19 weekly positivity trends, in terms of Granger-causality (by the GT keyword trend onto the COVID-19 weekly positivity trend; columns A and B) or Pearson correlation (columns C and D). For all the countries, the number of significant keywords was smaller in Granger-causality than in Pearson correlation (columns A vs. C, B vs. D), and the influence of multiple test adjustment (BH method) seemed to be larger in terms of Granger-causality (columns A to B) than in Pearson correlation (columns C to D). Specifically, the number of significant keywords identified by Granger-causality (with multiple tests adjusted: mean 11.7 ± 8.6 words) (Table 2, column B) decreased to approximately one-third when compared to those identified by unadjusted Pearson correlation (mean 32.1 ± 8.7 words) (Table 2, column C), especially in countries such as India, Japan, Singapore, and South Africa (outside Europe or North America). These results suggest that the current approach with appropriately adjusted Granger-causality analysis yields more stringent and statistically reliable results than the unadjusted Pearson correlation test, depending on the region.

**Table 2.**
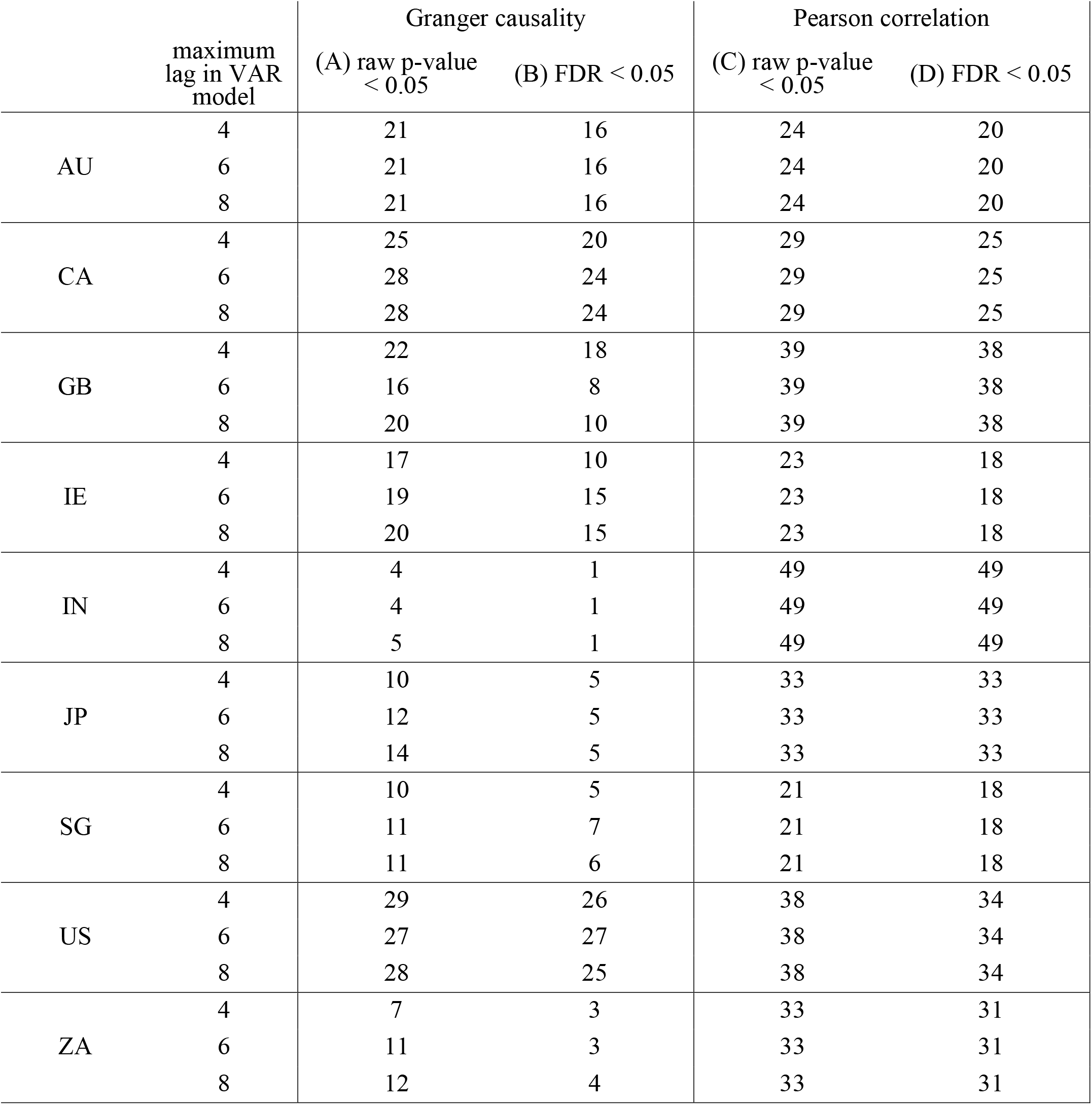
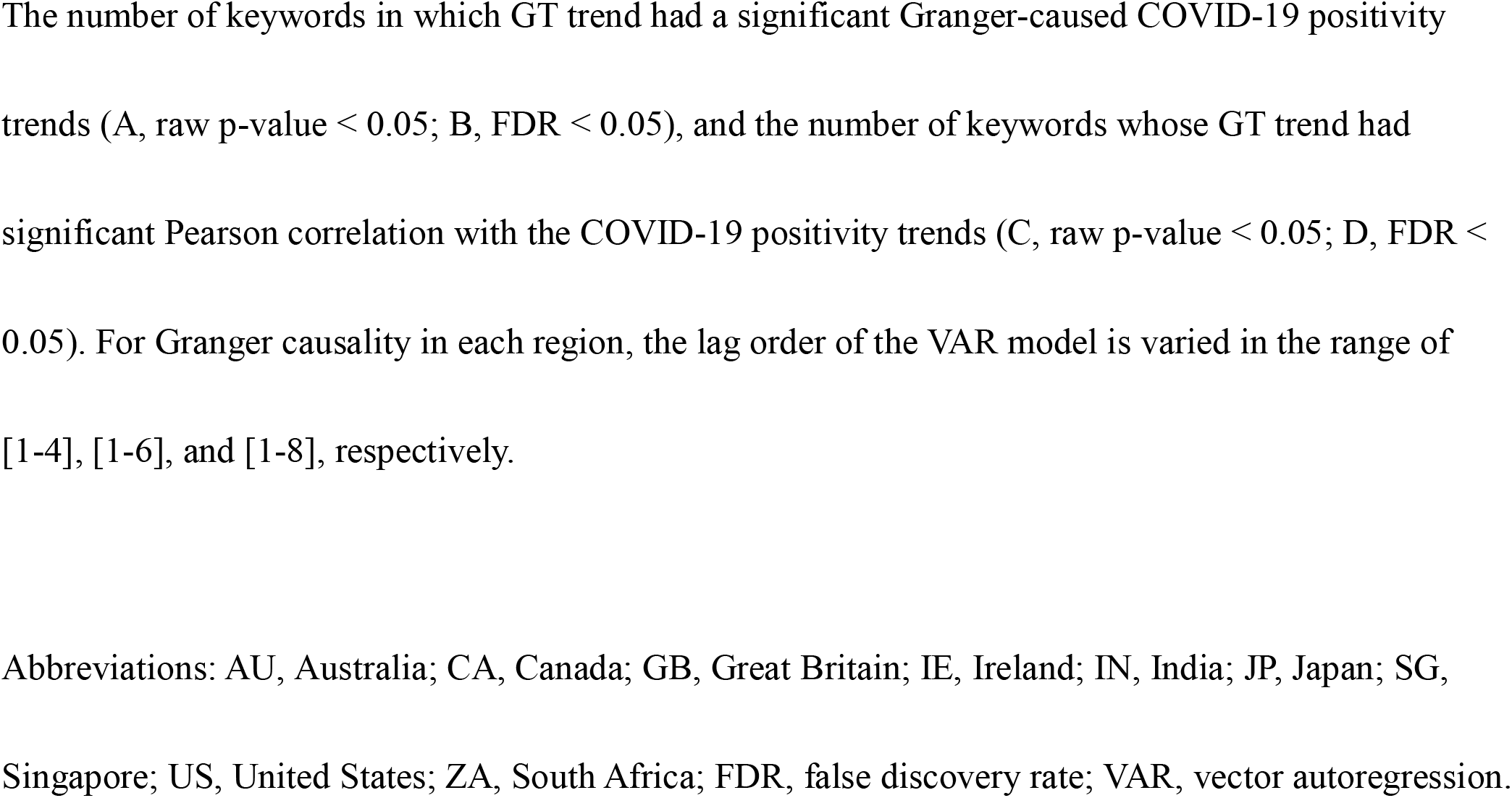
The number of significant keywords which have temporal association with the COVID-19 positivity trend.

|  | maximum<br>lag in VAR<br>model | Granger causality |  | Pearson correlation |  |
| --- | --- | --- | --- | --- | --- |
|  |  | (A) raw p-value<br>< 0.05 | (B) FDR < 0.05 | (C) raw p-value<br>< 0.05 | (D) FDR < 0.05 |
| AU | 4 | 21 | 16 | 24 | 20 |
|  | 6 | 21 | 16 | 24 | 20 |
|  | 8 | 21 | 16 | 24 | 20 |
| CA | 4 | 25 | 20 | 29 | 25 |
|  | 6 | 28 | 24 | 29 | 25 |
|  | 8 | 28 | 24 | 29 | 25 |
| GB | 4 | 22 | 18 | 39 | 38 |
|  | 6 | 16 | 8 | 39 | 38 |
|  | 8 | 20 | 10 | 39 | 38 |
| IE | 4 | 17 | 10 | 23 | 18 |
|  | 6 | 19 | 15 | 23 | 18 |
|  | 8 | 20 | 15 | 23 | 18 |
| IN | 4 | 4 | 1 | 49 | 49 |
|  | 6 | 4 | 1 | 49 | 49 |
|  | 8 | 5 | 1 | 49 | 49 |
| JP | 4 | 10 | 5 | 33 | 33 |
|  | 6 | 12 | 5 | 33 | 33 |
|  | 8 | 14 | 5 | 33 | 33 |
| SG | 4 | 10 | 5 | 21 | 18 |
|  | 6 | 11 | 7 | 21 | 18 |
|  | 8 | 11 | 6 | 21 | 18 |
| US | 4 | 29 | 26 | 38 | 34 |
|  | 6 | 27 | 27 | 38 | 34 |
|  | 8 | 28 | 25 | 38 | 34 |
| ZA | 4 | 7 | 3 | 33 | 31 |
|  | 6 | 11 | 3 | 33 | 31 |
|  | 8 | 12 | 4 | 33 | 31 |

The number of keywords in which GT trend had a significant Granger-caused COVID-19 positivity trends (A, raw p-value $< 0.05$ ; B, FDR $< 0.05$ ), and the number of keywords whose GT trend had significant Pearson correlation with the COVID-19 positivity trends (C, raw p-value $< 0.05$ ; D, FDR $< 0.05$ ). For Granger causality in each region, the lag order of the VAR model is varied in the range of [1-4], [1-6], and [1-8], respectively.
Abbreviations: AU, Australia; CA, Canada; GB, Great Britain; IE, Ireland; IN, India; JP, Japan; SG, Singapore; US, United States; ZA, South Africa; FDR, false discovery rate; VAR, vector autoregression.

The detailed results of the keywords that had significant Granger-causality (FDR < 0.05) to the weekly COVID-19 positivity trends are shown in Table 3 in decreasing order of identified frequency across the nine countries. Only keyword trends that had significant Granger-causality in four or more countries (out of the nine countries) are listed. The asterisk indicates that the keyword (in row) had significant Granger-causality in that country (in column). The lag order of the VAR model of each keyword is determined from the range of 1-4. The anosmia-related keyword “loss of smell” (or its corresponding Japanese keywords (S1 Table)) was identified in all nine countries, and the keyword “sense of smell” (or its corresponding Japanese keyword) was identified in five out of the nine countries.

**Table 3.** Top frequent keywords which significantly Granger-caused the COVID-19 positivity trends.

| keywords | total<br>frequency | regions |  |  |  |  |  |  |  |  |
| --- | --- | --- | --- | --- | --- | --- | --- | --- | --- | --- |
|  |  | AU | CA | GB | IE | IN | JP | SG | US | ZA |
| **loss of smell** | 9 | * | * | * | * | * | * | * | * | * |
| **sense of smell** | 5 | - | * | * | - | - | * | - | * | * |
| loss of taste | 5 | * | - | * | * | - | - | - | * | * |
| cough | 5 | * | * | * | * | - | - | - | * | - |
| runny nose | 5 | * | * | * | * | - | - | - | * | - |
| shortness of breath | 5 | * | * | * | - | - | - | * | * | - |
| sore | 5 | * | * | - | * | - | - | * | * | - |
| sore throat | 5 | * | * | * | * | - | - | - | * | - |
| stuffy nose | 5 | * | - | * | * | - | - | * | * | - |
| diarrhea | 4 | - | * | * | * | - | - | - | * | - |
| headache | 4 | * | * | * | - | - | - | - | * | - |
| pneumonia | 4 | * | * | * | - | - | - | - | * | - |

Detailed results of the keywords that have significant Granger-causality to the COVID-19 positivity trends, in the order of frequency across all the countries. Only keywords that were significant in 4 or more countries (out of 9) are shown. The asterisk indicates that the keyword (in row) had significant Granger-causality in that country (in column). The lag order of the VAR model of each keyword is determined from the range of 1-4.
Abbreviations: AU, Australia; CA, Canada; GB, Great Britain; IE, Ireland; IN, India; JP, Japan; SG,
Singapore; US, United States; ZA, South Africa; FDR, false discovery rate; VAR, vector autoregression.

Fig 3 visualizes GT search volume trends for “loss of smell” (or its corresponding Japanese word) for each country (dotted lines), which showed a clear temporal relationship with weekly COVID-19 positivity trends (solid lines). Other identified symptom keywords were as follows: “cough” (5/9 countries), “loss of taste” (5/9), “runny nose” (5/9), “stuffy nose” (5/9), “sore throat” (5/9), “sore” (5/9), “shortness of breath” (5/9), “diarrhea” (4/9), “headache” (4/9), and “pneumonia” (4/9). These are well-known symptoms of COVID-19 [2, 20] and partly overlap with the GT keywords reported to have significant associations with weekly COVID-19 case trends [5-7].

**Fig 3.**
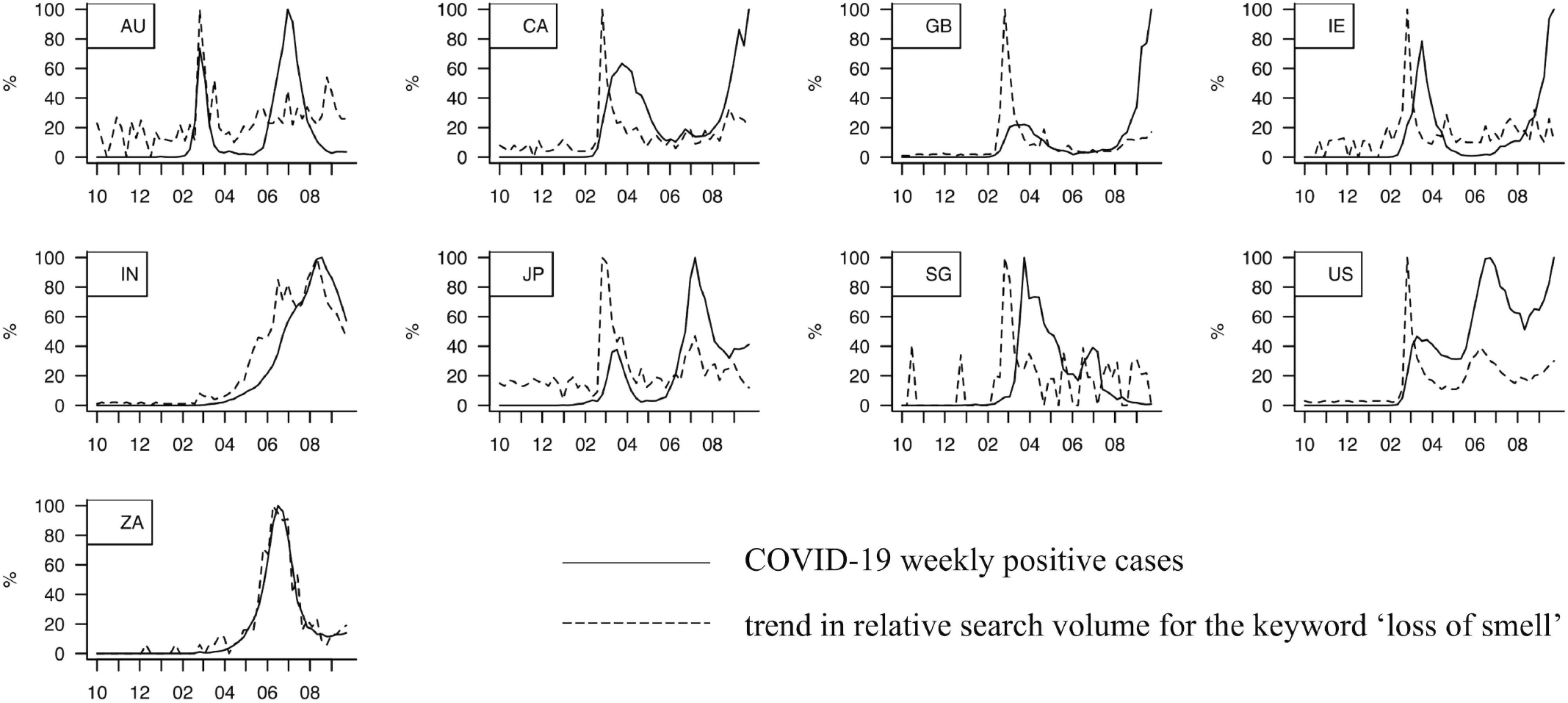
COVID-19 weekly positivity trends and the GT search volume trends for ‘loss of smell’ in each region. The relative GT search volume trends for the ‘loss of smell’ word (or its corresponding Japanese word) of each country (in dotted lines), which has clear temporal relationship with the COVID-19 positivity trends (in solid lines). X-axis in months since October 2019 to September 2020.

### Media coverage of keywords

The Japanese keywords corresponding to “loss of smell” and “sense of smell” were the only significant ones in Japan (Table 3, filled cells) and were also the most frequently identified keywords across the different countries, so we selected them to further assess the effect of media coverage trends on these keywords in the Japanese data. Fig 4 presents the temporal relationship between the weekly COVID-19 positivity trend (solid lines), the GT trend of the Japanese keywords (dotted lines) corresponding to (A) “loss of smell” or (B) “sense of smell,” and their media coverage trends (dashed lines). Apparently, in both keywords (A and B), the GT keyword trends were very similar to the trends in their media coverage. Notably, both the Granger-causality of the keywords “loss of smell” and “sense of smell” to the weekly COVID-19 positivity trend became non-significant when adjusted with their media coverage by partial Granger-causality analysis (*p = 0*.*257* and *p = 0*.*384*, respectively). These results suggest a relationship between weekly COVID-19 positivity trends and that the GT trends of anosmia-related keywords are highly confounded by their media coverage.

**Fig 4.**
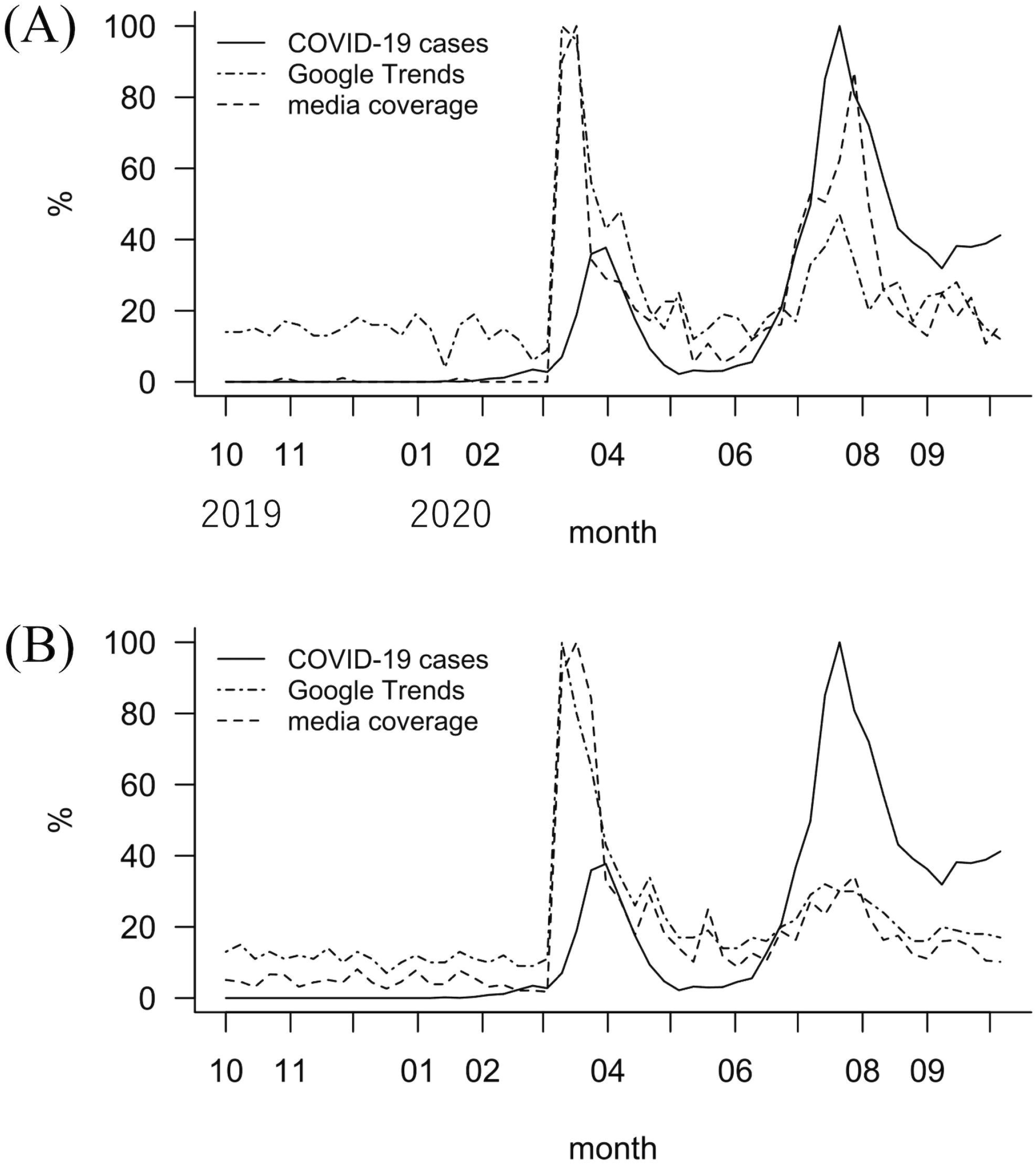
COVID-19 weekly positivity trends, Google Trends relative search volume trends for ‘loss of smell’ and ‘sense of smell’, and their media coverage trends in Japan. Temporal relationship between the COVID-19 positivity trend (in solid lines), the GT trend of the Japanese keywords (in dotted lines) corresponding to (A) ‘loss of smell’ or (B) ‘sense of smell’, and their media coverage trends (in dashed lines). Apparently, in both keywords (A and B), the GT keyword trends were very similar to the trends in their media coverage.

## Discussion

In summary, based on the potential analytical fallacies that are of concern in earlier GT studies, our current study aimed to identify symptom keywords in GT trends that could be used as a predictive measure for future weekly COVID-19 positivity trends by applying more statistically favorable methods. However, the current analysis showed that the number of search keywords that are truly associated with weekly COVID-19 positivity trends may be smaller than reported in earlier studies using a simple Pearson/Spearman correlation, of which the degree depends on the region. In addition, even the GT trends of most reliable anosmia-related keywords were actually a strong reflection of its media coverage (at least in Japan). These results suggest that many of the search keywords reported as candidate predictive measures in earlier GT studies may actually turn out to be false-positive. In other words, the potential candidate keywords listed in the earlier GT-based COVID-19 infodemiological studies are not always reliably usable as true predictive measures. We need to be careful when interpreting published study results as the utility of Google Trends for studying COVID-19 epidemiology may be more limited than previously expected.

The major strength of our study is its statistically favorable approach with a longer period of included observations. For example, our results evaluating the trend in media coverage of the “loss of smell” keyword is partly consistent with a few of the earlier studies [7, 9]. However, in previous studies, the potential effect of media coverage was not evaluated in a statistically favorable manner, and the association between GT trends and weekly COVID positivity trends had been evaluated in an inappropriate way (i.e., Pearson correlation). Moreover, earlier GT studies did not always examine many symptom keywords related to COVID-19 comprehensively as in our study, so that selection bias cannot be excluded. In contrast, our approach of narrowing down the candidate keywords to adjust for their media coverage was data-driven with a smaller risk of bias in keyword selection. In addition, because our study included a longer period of data (up until October 2020) than most of the earlier GT-based COVID-19 studies, which only included serial data within the first wave (e.g., up until July 2020 in the United States and Japan), lessons based on our results may have higher applicability to the second or later waves of weekly COVID-19 positivity trends.

Our study has some limitations. For example, in the VAR model, the effect of each variable is assumed to be fixed throughout the reviewed period, which may not always be true because the public interest and attitude toward COVID-19 could vary over time [21]. This can be suspected by the decreased peak of GT trend for the “COVID” keyword in the second wave (Fig 2, in Australia, Japan, and the United States). In future studies, state space modeling [22] to incorporate potentially time-varying effects may be useful to overcome the potential weakness of the VAR model, especially when the included period becomes so long. In addition, the keywords’ media coverage was adjusted only in Japanese regional data, which makes the obtained results slightly less generalizable to other countries. The *Nikkei telecom* we used for media review would not cover all potentially influencing media such as TV talk shows, or social media (e.g., *Twitter* [23] or *Instagram* [24]).

To conclude, our current results using a more statistically favorable approach suggest that many of the search keywords identified as candidate predictive measures in earlier GT studies have the potential risk of false positives, and that we need to be careful in interpreting the earlier GT-based COVID-19 study results.

## Data Availability

The data used in this study is retrieved from Google Trends (https://trends.google.co.jp/trends), a publicly-available source of search trafficking data.

## Acknowledgements

This work was supported by the Japan Society for the Promotion of Science (JSPS) KAKENHI Grant Numbers 20J11009 (K.S) and 20H03587 (A.I), and also supported by AMED under Grant Number 20dk0207048h0002.

