## Supplementary material for "Need of care in interpreting Google Trends-based COVID-19 infodemiological study results: potential risk of false-positivity": S1 Table

**S1 Table. English-Japanese corresponding table for symptom keywords searched**

| English keywords searched in Google Trends | | | Corresponding Japanese keywords searched | | | |
| --- | --- | --- | --- | --- | --- | --- |
| malaise |  |  | 倦怠感 | だるい |  |  |
| fatigue | tired |  | 疲労 |  |  |  |
| anorexia |  |  | 食欲不振 |  |  |  |
| diarrhea |  |  | 下痢 |  |  |  |
| constipation |  |  | 便秘 |  |  |  |
| abdominal pain | stomach ache |  | 腹痛 |  |  |  |
| nausea |  |  | 吐き気 |  |  |  |
| vomiting |  |  | 嘔吐 |  |  |  |
| chest pain |  |  | 胸痛 | 胸が痛い |  |  |
| dyspnea |  |  | 呼吸苦 | 呼吸困難 | 胸が苦しい |  |
| shortness of breath | short of breath |  | 息切れ |  |  |  |
| pneumonia |  |  | 肺炎 |  |  |  |
| cough |  |  | 咳 |  |  |  |
| sputum |  |  | 痰 |  |  |  |
| runny nose | nasal discharge |  | 鼻水 | 鼻汁 |  |  |
| stuffy nose |  |  | 鼻づまり |  |  |  |
| rhinitis |  |  | 鼻炎 |  |  |  |
| sneeze |  |  | くしゃみ |  |  |  |
| sore throat | throat pain |  | 喉が痛い | 咽頭痛 |  |  |
| fever |  |  | 発熱 | 熱 |  |  |
| chills |  |  | 悪寒 |  |  |  |
| cold |  |  | 寒気 |  |  |  |
| sense of smell |  |  | 嗅覚 |  |  |  |
| sense of taste |  |  | 味覚 |  |  |  |
| loss of smell | anosmia |  | 嗅覚異常 | 嗅覚障害 | においがしない | (無味無臭) |
| loss of taste | dysgeusia |  | 味覚異常 | 味覚障害 | 味がしない | (無味無臭) |
| hair loss | loss of hair |  | 抜け毛 | 毛が抜ける | 脱毛 |  |
| bald |  |  | はげ |  |  |  |
| myalgia | muscle pain |  | 筋肉痛 | 筋痛 |  |  |
| arthralgia | joint pain |  | 関節痛 |  |  |  |
| body aches |  |  | 全身の痛み |  |  |  |
| sore | pain |  | 痛み | 痛い |  |  |
| eye pain |  |  | 眼痛 |  |  |  |
| congestion |  |  | 充血 |  |  |  |
| headache |  |  | 頭痛 |  |  |  |
| vertigo | dizziness | dizzy | めまい |  |  |  |
| memory loss |  |  | 記憶障害 |  |  |  |
| confusion |  |  | 混乱 | 錯乱 |  |  |
| insomnia |  |  | 不眠 | 眠れない |  |  |
| anxiety |  |  | 不安 |  |  |  |
| numbness |  |  | しびれ | 痺れ |  |  |

The corresponding English-Japanese table for the searched symptom keywords: 54 English keywords (left-sided 3 columns) were used for search in 8 English-speaking country regions and the corresponding 60 Japanese keywords (right-sided 3 columns) were used for search in the Japan region. Words listed in the same row roughly belong to the similar symptom category.
